# Probability that an infection like Covid-19 stops without reaching herd immunity, calculated with a stochastic agent-based model

**DOI:** 10.1101/2020.12.27.20248905

**Authors:** Manfred Eissler

## Abstract

The spread of an infection is simulated with a stochastic agent-based model. In a certain range of R0 values, the infection either rapidly comes to halt or a large proportion of the population is infected until herd immunity is achieved. Which of these two possibilities actually occurs is random. The probability of each case is determined ‘quasi-empirically’. This stochastic phenomenon may explain unexpected infection trajectories.

## Background

Agent-based infection models are excellent at simulating and visualizing the course of an infection. One advantage over deterministic models calculated with differential equations is that the progression over time can be modelled stochastically, and interventions can be incorporated into the sequence. This allows for example the formation of a second wave to be simulated with straightforward models. More complex agent-based models use real data for the simulation of infections [1],[2],[3].

The results of stochastic processes are usually random variables. In the model considered here, the total number of individuals who went through the infection once the infection event has subsided is a random variable that fluctuates around a mean value. These values are determined “empirically” with a simple model by performing multiple runs with the same parameters in each case.

## Method

The spread of an infection is simulated with an agent-based SIR-model. The agents (= individuals) are points (identified by integer coordinates) in a rectangular coordinate system. Initially, every agent is susceptible to infection. The process begins with one infected individual (at the origin). Each infected individual can infect the directly adjacent individuals to the left, right, above, and below. Thus, the first infected individual can infect up to four other individuals immediately adjacent to it. A predetermined number of these four individuals are randomly selected, e.g. two new individuals, who are then considered to be infected. This allows the basic reproduction rate R0 to be specified. This process is iterated after defining the recovery rate β, which describes how long an infected individual remains sick and able to infect others. In the model considered here, the duration of the disease and the infectious period are taken to be equal. At the end of the period, the infected individual joins the group of restored individuals. In a simple model with no mobility of infected individuals, the spread is essentially centrifugal, ending once a certain percentage of individuals have been infected and ultimately join the restored group. This proportion depends on R0 and corresponds to the phenomenon of herd immunity.

In a more realistic variant of the model, the infected individuals are permitted to move around. If the number of newly infected individuals exceeds a specified value in a given iteration, a configurable number of infected individuals are randomly distributed throughout the entire population. This creates new infection cells that form into new clusters. By switching the mobility on and off, the formation of a second wave can be simulated.

In another variant, the initial situation can be changed in such a way that a certain percentage of the population is immune from the start, e.g. due to vaccination or immunity of a certain percentage from a previous wave. Since the infection unfolds stochastically in these simulations, the final state is a random variable whose values may differ between runs, and for which a mean value can be determined “empirically” by performing multiple runs. This was performed for different values of R0, different vaccination rates and ‘pre-immunization measures’, and different population sizes.

The simulation was carried out using the program “Wolfram Mathematica Version 11.3”.

## Results

With a population size of n0 = 10,000 without any initial immunity, the maximum number of infected individuals when the infection ends, i.e. when herd immunity is achieved, was determined by performing multiple runs in each case for different R0 values. In a specific range of R0 values, an unexpected phenomenon occurs: a dichotomy in the final values (Fig. 1). For R0 values ranging from to 2.0, a total of 20 runs were performed in each case. The runs are numbered from 1 to 20. At small R0 values (here, R0 = 1.2), only a few individuals are infected, and the infection comes to a halt after only a few time steps in each of the 20 runs. As R0 increases, significantly higher values of the total number of infected individuals occur with increasing frequency. Since these values correspond to herd immunity, they increase with R0, as expected. There is a clear gap between the large and small values – the results do not form a continuum. The values are either relatively close to a high value or very densely located around a low value that is close to zero.

**Fig 1.**
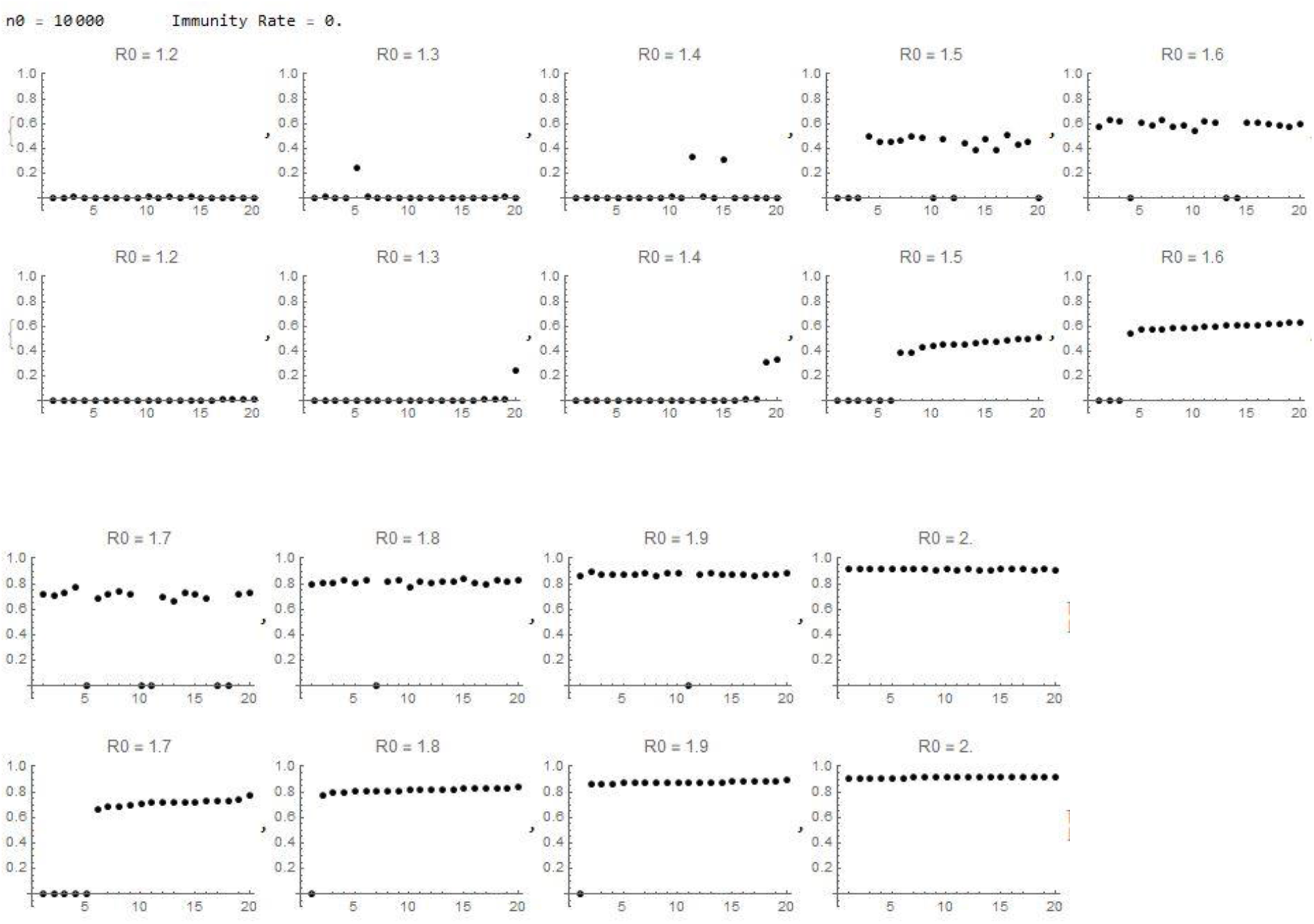
Results for the number of infected individuals for 20 runs with different R0 values. The top row shows the proportion of infected individuals in each run of the simulation, the bottom row the same values, ordered by size.

In Fig. 2, the values are summarized in a graph. Each value is random. By performing a large number of runs (here, 20 runs), the mean values and ‘empirical’ probabilities of the occurrence of large and small values can be determined (Fig. 3).

**Fig 2.**
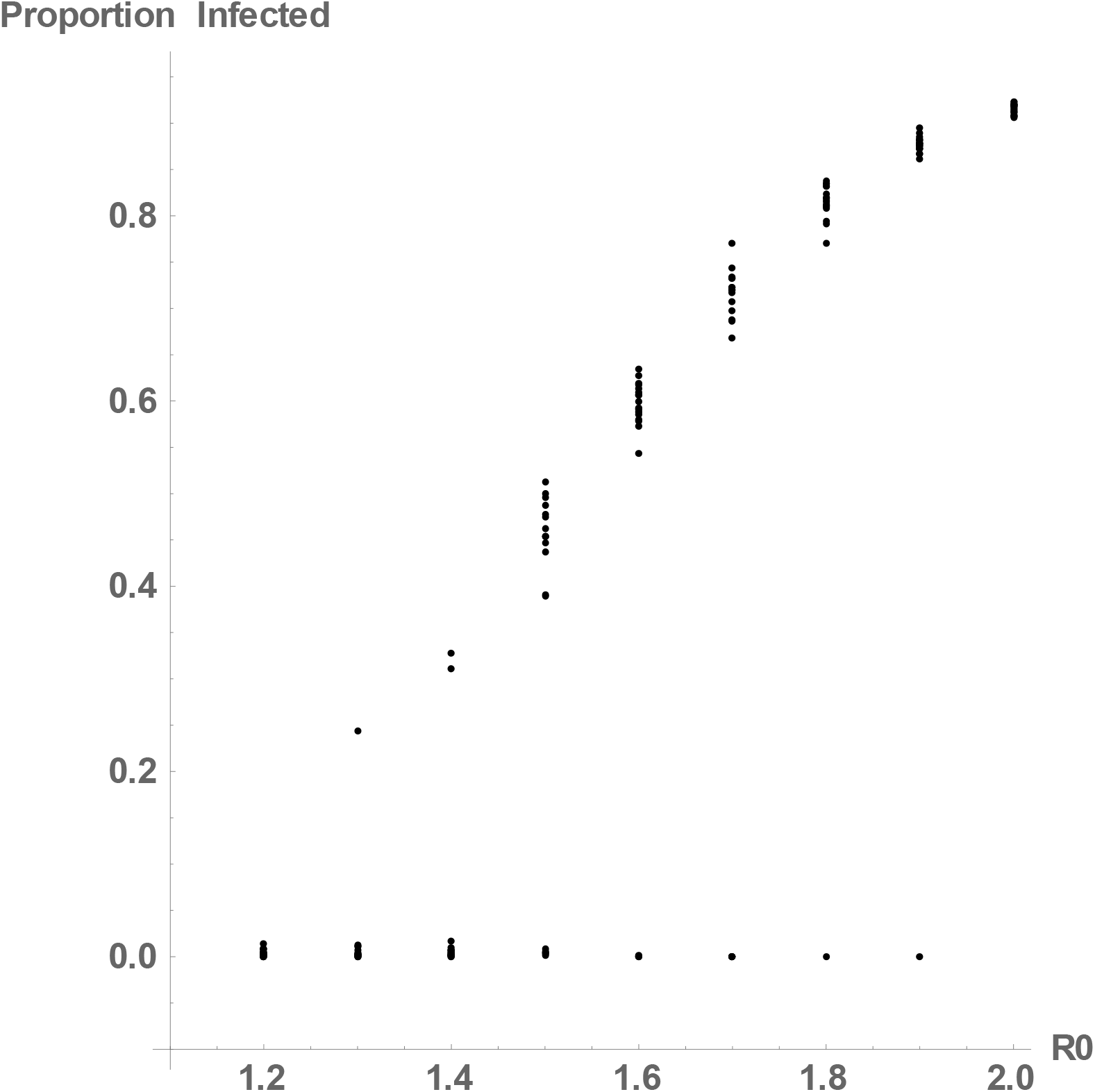
Results for the number of infected individuals for various R0 values. For each R0, 20 points are plotted, some of which are very close to one another. To make the graph easier to read, the y-axis was shifted by -0.1.

**Fig 3.**
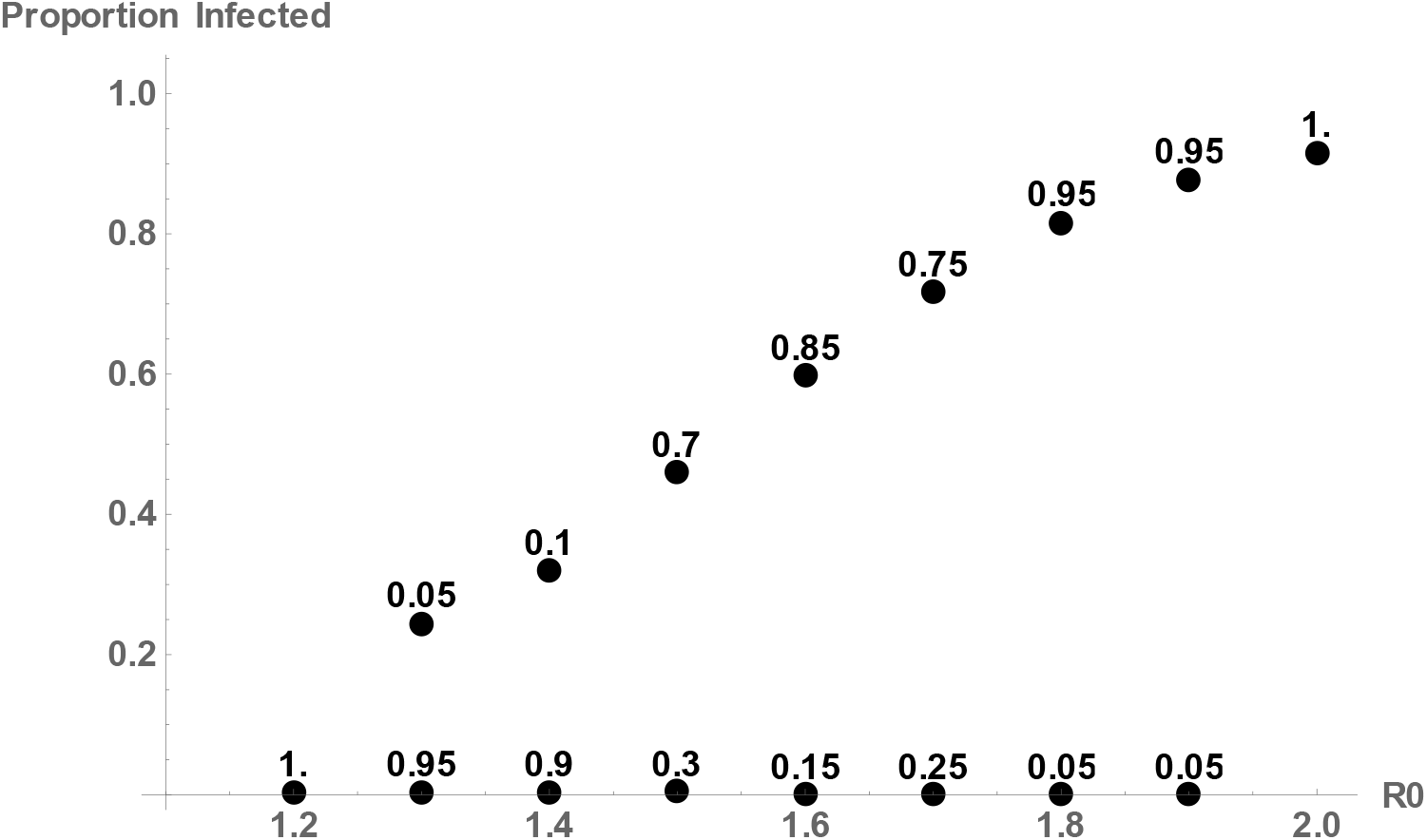
The means of the large and small values and the probability with which each case occurs. (n0 = 10000, number of runs per R0 = 20, initial immunity rate = 0)

If the population is ‘pre-immunized’, either by vaccination or following a first wave of infection, the probability gap shifts to higher R0 values. Fig. 4 shows the results for an immunization rate of 0.1, and Fig. 5 shows the results for an immunization rate of 0.3.

**Fig 4.**
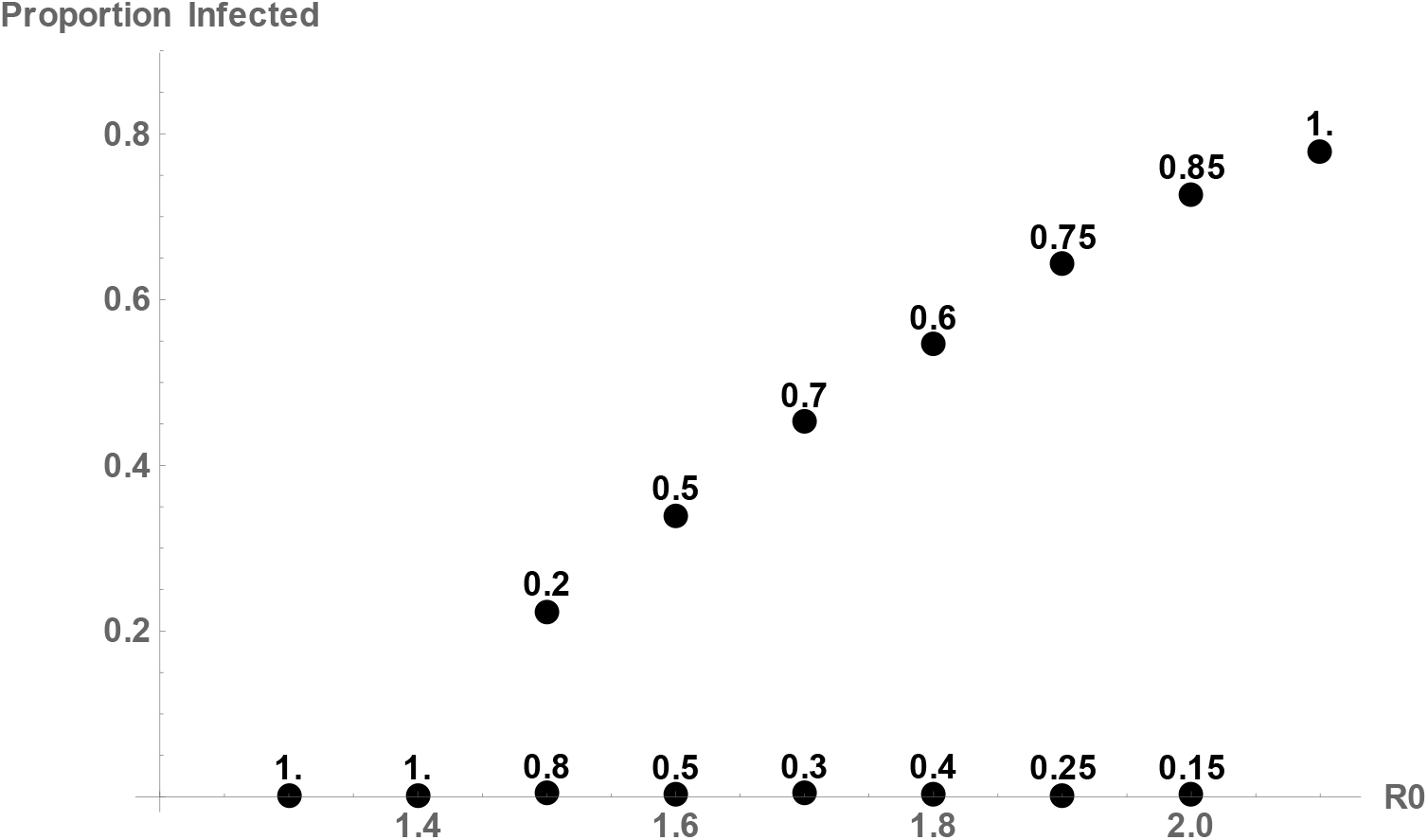
The means of the large and small values and the probability with which each case occurs. (n0 = 10000, number of runs per R0 = 20, initial immunity rate = 0.1)

**Fig 5.**
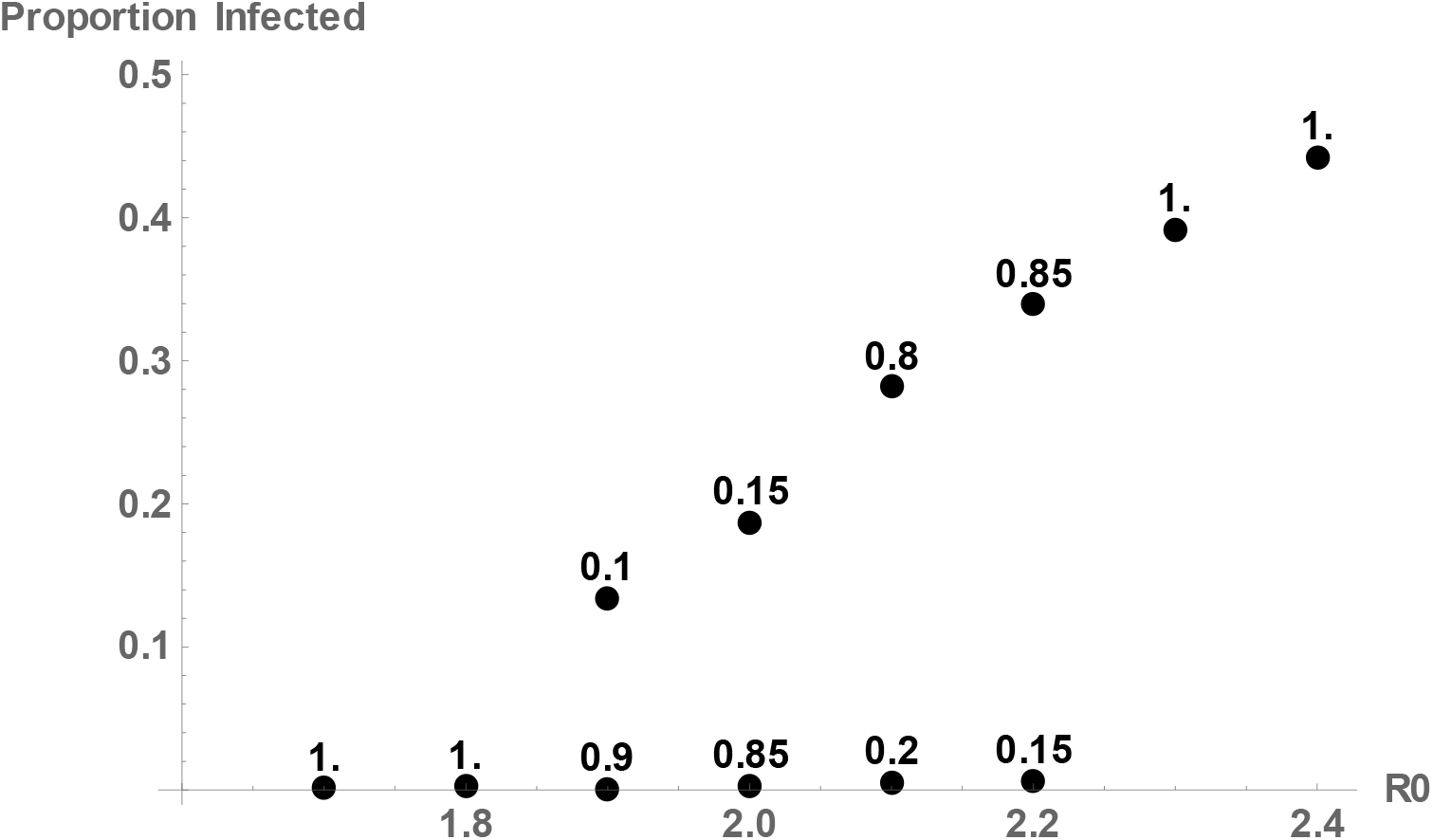
The means of the large and small values and the probability with which each case occurs. (n0 = 10000, number of runs per R0 = 20, initial immunity rate = 0.3)

The effect is less clear with smaller populations, but it becomes more pronounced as the population increases, i.e. the statistical fluctuations around the mean become relatively smaller.

## Discussion

Since the infection unfolds stochastically in these agent-based simulations, the final state is a random variable whose value may differ between runs, and for which a mean value can be determined ‘empirically’ by performing multiple runs. In this regard the stochastic agent-based model differs from deterministic simulations. As such, a stochastic model likely gives a more accurate description of a true infection, which can certainly be described as a stochastic process.

The more complex agent-based models by Hunter et al [2], which simulate measles outbreaks in Irish cities with realistic data, show qualitatively comparable results for the random variables ‘mean of infected people’ and ‘probability of an outbreak’. However, a direct comparison is not possible because R0 values from 15 to 150 were used for the simulation. Here the low range of R0 is examined, which could be of relevance to infections such as influenza or Covid-19.

The model used here is a relatively simple stochastic agent-based model. Whether the observed probability gap plays an important role in the course of real infections must be answered by future studies. If the findings are transferable, it could mean that even a relatively low vaccination quota might cause an infection to disappear from a population with a certain probability if the corresponding R0 values are reached and maintained, without requiring herd immunity to be achieved. If so, the gap could be described as a ‘gap of good hope’. However, it would not imply that infection can no longer occur. Isolated clusters would still remain possible. The infection will only be extinguished if infected individuals ‘from outside’ can be consistently prevented from entering the population, which remains partially susceptible.

The next step is to calculate a mathematical and probabilistic basis for the model presented here. It should also be investigated whether the same effect continues to manifest in more complex agent-based models. The question of whether there are indications of such an effect in real infection processes is of course of great interest. This stochastic phenomenon might offer an explanation for the occasionally surprising course of infections in relatively well-isolated populations, regions or cities.

## Data Availability

all data are available

https://www.magentacloud.de/share/4t360-uwvv#$/

